# How do Emergency Departments respond to ambulance pre-alert calls? A qualitative exploration of the management of pre-alert in six UK Emergency Departments

**DOI:** 10.1101/2023.12.21.23300123

**Authors:** Jaqui Long, Fiona C. Sampson, Joanne Coster, Rachel O’Hara, Fiona B Bell, Steve Goodacre

## Abstract

**Background:** Whilst pre-alerts have been shown to improve outcomes for some patients requiring immediate time-critical treatment (e.g. stroke), little is known about their usefulness for other patients and what processes are used by Emergency Department (EDs) to respond to them. In the current context of high demand, it is important to understand how pre-alerts affect EDs.

**Methods:** We undertook non-participation observation (162 hours, 143 pre-alerts) and semi-structured interviews (40 staff) within six UK Emergency Departments (EDs), between August 2022-April 2023. Interview transcripts and observation notes were imported into NVivo™ and analysed using a thematic approach.

**Results:** Pre-alert calls involved significant time and resources for ED staff but enabled staff to prepare for patient’s arrival both practically and psychologically, particularly when demand was high. High demand created additional pre-alerts due to advice or ‘heads up’ calls from ambulance clinicians concerned about handover delay. Despite some pre-alert fatigue regarding patients who did not always require a special response (e.g. sepsis), ED clinicians prioritised and valued pre-alerts, perceiving higher risks from under-alerting than over-alerting. Variation in ED processes for a) senior clinical review of pre-alerted patients not brought into resus and b) receiving, documenting and informing others of pre-alerts resulted in inconsistent response to ambulance clinicians. ED response (where the patient should be taken) largely reflected resources available (beds, staffing, acuity of other patients) rather than appropriateness of the pre-alert.

**Implications:** In the context of high demand, much variation in response to pre-alerts is outside ED staff’s control. There is potential for EDs to increase consistency in reviewing how pre-alert calls are answered, what information is documented and how this is communicated to others, including when they are not accepted into resus. Improved communication between ambulance and ED services may help avoid tension caused by different perceptions or understandings of pre-alerts.

**What is known on this topic?:**

- Ambulance pre-alerts can help Emergency Department staff to prepare for a patient’s arrival and can lead to improved outcomes for patients requiring immediate senior review upon arrival.
- Research about pre-alert practice focuses on outcomes for patients who have been pre-alerted but there is a lack of evidence about the effect of pre-alerts on ED staff and ED patient management.

**What this study adds:**

- Variation in ED processes, layout and capacity led to different ED responses to pre-alert calls, particularly for patients who were not brought into resus.
- ED response is primarily dependent upon resources available at the time of the call and assessment of the need for active treatment. Pre-alerted patients who may be suitable for a resus bay may be seen in another area of the ED when the ED is crowded or has higher priority patients
- Pre-alerts used ED resources but were valued in terms of enabling both practical and psychological preparedness.

**How this study might affect research, practice or policy:**

- Standardisation of processes for improving flow and assessing high risk patients may help reduce variation in ED management and provide more consistent support for ambulance clinicians
- Understanding that EDs may not be able to provide an expected or consistent response to pre-alert calls is important for ambulance clinicians assessing their own pre-alert practice.

## Introduction

Ambulance clinicians may use a pre-alert call to the receiving hospital when they consider a patient requires a different or special response (1). Evidence suggests that pre-alerts lead to improved patient outcomes for certain conditions where patient pathways indicate the need for a specific and timely response (e.g. initiation of treatment, preparation of trauma team personnel).(2-7) Guidelines for management of patients with major trauma, sepsis, stroke and cardiac arrest all include recommendations for pre-alert use. (8-10) In the UK, joint guidance from RCEM/AACE also recommends pre-alerting for a range of other conditions or physiological criteria although there is significant variation in guidance recommendations for pre-alert and of pre-alert rates for different conditions across different ambulance services. (11, 12)

In the current context of increased ED crowding and ambulance handover times, pre-alerts can be key to ensuring critically ill patients can bypass ambulance queues and receive timely care. (1, 13) However, reducing thresholds for pre-alerting due to concerns about handover delays may lead to inappropriate use of pre-alerts, with resources being diverted from other urgent patient care and increasing the likelihood of pre-alert fatigue. (14-18).

Despite recognition of the need to balance the use of pre-alerts to ensure patient benefit without inappropriate deployment of resources (9, 19) there is a lack of research understanding how pre-alerts influence patient care in the ED, or the potential impact of increasing/changing use of pre-alerts on ED staff and patients. Concerns about pre-alert fatigue are primarily anecdotal and the impact of pre-alerts on ED staff is unknown. As part of a mixed methods study exploring the impact of pre-alerts on ED and ambulance staff and patients, we undertook qualitative research to explore how pre-alerts influence management and care of ED patients, including potential benefits and unintended consequences.

## Methods

We used a qualitative design, incorporating semi-structured interviews and non-participant observation.

### Patient and Public Involvement (PPI)

The study PPI group, which includes people with lived experience of pre-alerts, met regularly through the study. The PPI group reviewed and discussed the interview schedules and the emerging findings from the interviews. The group’s experience of pre-alerts also helped to inform observation practice.

### Context and Sampling strategy

Six ED sites were identified by selecting one Major Trauma Centre (MTC) and one Trauma Unit (TU) within each of three ambulance services, focusing on those with high numbers of pre-alerts to ensure that sufficient pre-alert activity could be observed during the researcher visits. Sites were selected to cover diverse populations in terms of deprivation, rural/urban mix and diverse ethnic populations.

Non-participant observations and informal conversations with ED staff were undertaken at each site. We recruited ED staff for interviews primarily through direct invitation during observations, with local investigators also asked to invite staff with particular roles (e.g. clinical director). We aimed to recruit a range of different roles at each site, including senior and junior medical and nursing staff as well as other roles identified as important at individual sites during the fieldwork.

### Data collection methods

Observation and interviews were undertaken principally by 2 researchers (JL & JC) with some dual observations where departments were particularly large or busy and required observation in multiple areas. Researchers were principally based near to the pre-alert phone (usually in resus) in order to be able to observe and record the ED response to calls, but also observed throughout the ED and the ambulance waiting areas. Staff were made aware of the presence of researchers and given the option to opt out of being observed. Further details of observations are available in supplementary file (1).

### Data collection instruments, technologies and processing

We developed interview topic guides in collaboration with our project management and PPI group. Topic guides were followed flexibly. Observation guides were developed and refined after initial visits, including a form to record key details of individual pre-alert calls (not recording any patient data). Interviews were conducted online or by phone and recorded using encrypted dictaphones and transcribed verbatim. Data was stored in a restricted area of the university secure filestore, accessible only by the research team at University of Sheffield. All participants were allocated a unique code, which was used within data excerpts. Transcripts were not made openly available due to concerns about anonymity. All fieldwork data (interview transcripts and observation notes) were loaded into NVivo. (20)

### Researcher characteristics and reflexivity

The researchers involved in the data collection were experienced researchers working in health services research with social science/psychology background but not clinically trained. Two of the researchers (FS and ROH) had prior experience of undertaking non-participant observation in emergency settings, while the researchers involved in fieldwork (JL/ JC) had no prior experience and thus fewer pre-conceptions about how emergency services worked. Observation notes were written up in detail shortly after the observation took place to incorporate researcher reflections and interpretation of events.

### Data analysis

Data was analysed using a thematic approach according to the principles of Braun & Clark. (21) Data familiarisation involved ROH, JL, JC and FS reading a subset of the interviews and developing initial themes and an initial coding framework. Data was coded independently, undertaken initially by ROH (who had not undertaken any fieldwork) and JL (who had done the majority of data collection). Coding was discussed and refined within the wider group on a weekly basis in order to refine analysis. Codes and changes to coding frameworks were documented at each stage. Code summaries were developed and cross-cutting themes identified after discussion between the group.

### Techniques to enhance trustworthiness

Researchers clarified points and summarised findings during interviews to ensure a shared understanding of the data. Researcher triangulation within both the data collection and analysis phases helped improve trustworthiness of analysis. Results were presented to PPI at an online workshop and their views of the findings and which findings were most important to PPI contributed to a wider stakeholder workshop incorporating research participants and key stakeholders from ambulance service and ED national bodies on how to use the findings to improve practice.

### Findings

We undertook interviews with 40 ED clinicians (see table 1 for details).

**Table 1:** Characteristics of ED clinician interview participants.

|  |  |  |  |
| --- | --- | --- | --- |
| **ED** | Major Trauma Centre | Site A | 7 |
|  |  | Site D | 8 |
|  |  | Site E | 6 |
|  | Trauma Unit | Site B | 4 |
|  |  | Site C | 10 |
|  |  | Site F | 5 |
| **Role** | Consultant |  | 16 |
|  | Registrar |  | 7 |
|  | Junior doctor |  | 2 |
|  | Senior nurse |  | 10 |
|  | Nurse |  | 2 |
|  | Practitioner |  | 3 |
| **Years in role** | <1 year |  | 8 |
|  | 1-5 years |  | 20 |
|  | 6-10 years |  | 6 |
|  | >10 years |  | 6 |
| **Gender** | Female |  | 23 |
|  | Male |  | 17 |
| **Ethnicity** | White British |  | 33 |
|  | British Asian |  | 4 |
|  | Not reported |  | 3 |

We undertook 25 sessions of observations across six EDs, completing a total of 162 hours (or 123 hours of actual observed time) and observing 143 pre-alert calls (see table 2). Sessions ranged from 2.25-7.25 hours, average 5 hours. Further details of pre-alerts are available in supplementary material.

**Table 2:** Details of pre-alerts observed.

| Site | No. alerts observed | Type of alert | No. hours observed | No. Resus/ high care | Front door<sup>+</sup> | Senior clinician triage | No other |
| --- | --- | --- | --- | --- | --- | --- | --- |
| A* (MTC) | 26 | 21 medical, 5 trauma | 31.5, 5 days | 15 | 11 | 0 | 0 |
| B (TU) | 6 | 6 medical, 0 trauma | 14, 3 days | 5 | 0 | 1 | 0 |
| C (TU) | 34 | 27 medical, 7 trauma | 28, 6 days | 16 | 0 | 11 | 7 |
| D* (MTC) | 28 | 26 medical**, 2 trauma | 35.5, 4 days | 20 | 0 | 5 | 3 |
| E* (MTC) | 24 | 19 medical, 5 trauma | 25, 3 days | 18 | 0 | 1 | 5 |
| F* (TU) | 25 | 25 medical, 0 trauma | 28, 4 days | 16 | 8 | 0 | 1 |

\* includes some hours double observation (A=8hrs; D=18hrs; E=8hrs; F=5hrs)
\*\* one alert classed as both medical and trauma
+front door is the term used for the main ambulance entrance (e.g. pitstop)

### Description of cases

A brief description of each site and its processes for managing pre-alerts is in Table 3.

| Site code, type of site (MTC/TU) | Resus provision.<br>Alternative options if not accepted to resus. | Access to resus for ambulance crews | Location of red phone(s). Who answers? | Pre-alert documentation.<br>How information about prealert is communicated to others, including use of documentation | Who is involved in prealert decision-making?<br>Key staff involved in management of alerts. |
| --- | --- | --- | --- | --- | --- |
| A – MTC | 8 resus beds, with trauma/ high acuity bays nearest ambulance entrance.<br>Alternative: initial assessment area | Direct from outside or from ‘front door’ following assessment. Crews only bring direct to resus if this has been agreed on the phone | In resus, but audible throughout the department. Policy is to be answered by consultant, but answered by whoever is nearest. | One form for all calls. Form either goes by patient’s bed in resus or is taken to ‘front door’ for the receiving nurse/staff. Other relevant staff informed verbally by call-taker. | Decision made by person answering, with input if needed. Consultant and NIC assigned specifically to resus. |
| B – TU | 5 resus beds, with trauma/high acuity bay nearest ambulance entrance.<br>Alternatives: a) 2 high dependency beds; b) initial assessment area | Either direct from outside, or from assessment area. Crews only bring direct to resus if this has been agreed on the phone? | In majors, on main desk where doctors are sitting. Answered by doctor generally, as tend to be nearest person to the phone. | Separate trauma and medical forms. Form taken to resus, high dependency cubicle, or left by ambulance handover bays for nurse receiving ambulance crew. Other relevant staff informed verbally by call-taker. | Decision made by person answering, with input if needed. Consultant has oversight of resus, high dependency and majors, |
| C – TU | 4 resus beds<br>Alternatives: a) 4 beds with higher staff/ patient ratio; initial assessment | Off main corridor into the department only. Crews only bring direct to resus if this has been agreed on the phone. No other access route. | In majors, at NIC desk. Not audible in other areas of department. Answered by NIC when possible, by whoever is | One form for all calls. Form taken to resus or high care area, or given to assessment nurse in ‘front door’ area receiving ambulance crew. Other relevant staff, including HALO, | Most decisions made by NIC, with consultant/medical input when needed. Consultant manages resus and high care, others in majors. |

|  | area or assessment on ambulance |  | nearest if not. | informed verbally by call-taker. |  |
| --- | --- | --- | --- | --- | --- |
| D – MTC | 5 resus beds, with trauma/high acuity bays nearest ambulance entrance<br>Alternatives: a) 6 bed rapid assessment & treatment area; b) direct to majors | Off the main corridor from the front door and majors area. Crews only bring to resus if agreed on the phone. No other access route. | 2 phones in resus at staff desk; a third phone rings in majors if other two lines engaged.<br>Answered by whoever is nearest who feels confident to do so, often ODP. | Separate trauma and medical forms in folders.<br>Forms generally remain in folders. Relevant staff, including rapid assessment area staff, receiving nurse & HALO, informed verbally and/or through a 'bleep' system via main switchboard. | Decision mostly made by person answering, with additional input if needed. Consultant informed of/approves all decisions re patients NOT accepted to resus.<br>Consultant cover from majors, variably in resus much of time |
| E – MTC | 9 resus beds, some of which can be divided, with trauma/high acuity bays nearest ambulance entrance.<br>Alternatives: initial assessment area | Immediately off the corridor by the ambulance entrance. Crews can drop in and ask about patients they are concerned about but haven't alerted.<br>Also an entrance from 'front door'/majors | In resus, at NIC desk.<br>Answered by NIC mostly, but whoever is nearest. | One form for all.<br>Forms either goes by patient's bed in resus or may be taken to 'front door' area but not consistently – some are left in a pile by the red phone. Other relevant staff in resus informed verbally. | Decision mostly made by person answering, with additional input when needed.<br>Consultant and NIC assigned specifically to resus.<br>Consultant variably involved in decision-making. |
| F – TU | 7 resus beds, with trauma bay nearest ambulance entrance.<br>Alternative: initial assessment area | Through 'front door' area. Crews can't access without going through pit stop. Crews only bring to resus if agreed on the phone. | In resus, at staff desk. Bell also rings in majors, making them aware of the call.<br>Answered by whoever is nearest, generally NIC or more senior doctor. | One form for all.<br>Forms either go by patient's bed in resus or are taken to 'front door' and handed to NIC or doctor. Other relevant staff informed verbally. | Decision mostly made by person answering, with additional input when needed.<br>Consultant cover from majors. |

\* The term 'front door' is used to describe all department's initial assessment and treatment area i.e. where those patients not being taken to resus etc are received.

**Table 4:** Outline description of observation sites and pre-alert processes.

| Site | Brief description |
| --- | --- |
| A<br>MTC | Department, including resus, often full, and ambulances frequently queuing outside. Some assessment of patients on ambulances at particularly busy times. Hospital Ambulance Liaison Officer (HALO) also present at these times to help prioritise queues, support ambulance crews. Resus has an allocated consultant, nurse in charge and other medical and nursing staff. Pre-alert phone rings very loudly. Information regarding pre-alerts not accepted into resus generally reliably conveyed to staff in 'front door' area. |
| B<br>TU | Relatively spacious, with a large number of computer terminals. pre-alerts a much smaller part of the workload, occurring much less frequently than at other sites during observations. Resus area separate from the rest of the ED, and only staffed when patients were there – it was rarely full. Ambulance crews rarely queue for any length of time for assessment, even when not pre-alerted. |
| C<br>TU | Very overcrowded, with patients frequently assessed and managed in corridor and on ambulances. Department generally full, with ambulances often queueing for long periods. pre-alert phone inaudible at any distance a significant issue. pre-alerts often required significant 'reshuffling' of multiple patients between areas and communication with numbers of staff to make space – nurse in charge has key role and answers the phone most of the time to facilitate. HALO paramedic often on site to facilitate management of queues, sometimes providing additional information on incoming patients. ED staff had access to ambulance crews' electronic patient records before arrival. |
| D<br>MTC | Core resus staffing included specialist practitioners who had key role in answering phone and treating patients. 3 phone lines for pre-alerts. Details of alerts not accepted into resus conveyed verbally, but generally reliably. 'Bleep' system via switchboard used to notify key staff of incoming alerts. pre-alert paperwork not linked to patient notes. HALO generally present and with key role in facilitating communication. Department generally full, with ambulances often queueing. Rapid assessment area provided an intermediate level of response for some pre-alerts. Variable level of consultant input into resus, depending on individuals and demand. ED staff had access to ambulance crews' electronic patient records before arrival. |
| E<br>MTC | Large resus area, with capacity for further sub-division at busy times. Much smaller initial assessment area for patients not accepted into resus, but crews sometimes called into resus on arrival with non-alerted patients for a quick assessment, and this was accepted by resus staff. Information regarding pre-alerts not accepted into resus not consistently recorded or passed to 'front door' staff. Patients generally not held on ambulances but booked in and then queuing on trolleys along corridors, often for long periods. Resus has an allocated consultant, nurse in charge and other medical and nursing staff. Consultant input into decisions varied depending on the individuals and demand. |
| F<br>TU | Resus largely managed by nurse in charge and 'junior' doctors, including experienced registrars – consultant based in majors and provided input depended on level of experience of resus staff and demand. Alert phone triggers bell in majors department but does not prompt any specific response. Initial assessment area very busy, with frequent movement through to majors area as beds became free. Some tension observed when pre-alerts not accepted into resus and passed to 'front door' staff – concern re risk, ability to manage. Very busy department, with ambulances often queuing, though generally not for long periods. |

We identified variation at each stage of the pre-alert response, including how pre-alert calls were received in the department, who took the call, how calls were documented, ED response and processes for managing and communicating the response to other staff.

### 1. ED response (i.e. where the patient should be taken) reflected resources available more than appropriateness of pre-alert

During fieldwork EDs often had high levels of crowding and frequent ambulance queues of ambulances. ED response (i.e. where they told the ambulance clinician to bring the patient) varied depending on the resources available at the time of call and did not always reflect whether a resus bed would be appropriate for the patient, but whether the patient was perceived to be more ill than existing patients. When departments were busy, this involved balancing who was least sick (rather than most well) and/or directing patients who at a quieter time may have been offered a resus bed to a different area of the ED. Pre-alert demand was often high and repeated calls in quick succession created additional pressures, particularly when space in other areas was limited.

*[SiteD MTC Obs 4a]: Clinician in charge arrives - “I hate shifts like this, who’s coming out?” and says “they’re trying to work out who’s the least worst, not who’s well enough to move out, but who’s the least likely to arrest on me if I move them out there.”*
*[SiteD MTC Obs 1a]: Conversation between the nurse in charge and [role] about the two pre-alerts. ‘What’s yours? Is yours more important than mine? We’ve only got one space. This one might be able to go to [senior clinician triage]’. The [flow manager] goes to check with majors about where each patient should go*.
*Another [Flow manager] announces “medical alert to resus in 5 minutes”. Staff are getting the bed ready. [Flow manager] tells me they’re not sure what’s happening with the other pre-alert She thinks it’s going to [senior clinician triage]*.

ED clinicians considered the need for active treatment (rather than watchful waiting) and the ability to manage certain patients or conditions outside resus, despite considering them valid pre-alerts. Sepsis in particular was recognised as a frequently alerted condition but often manageable outside of resus. ED clinicians talked about ‘pre-alert fatigue’ in relation to sepsis (and other) alerts, particularly where ambulance clinician protocols required them to pre-alert (courtesy calls) but ED staff did not always feel that a resus bed was required.

*[ED38]: Most of our alerts are sick medical cases, and sick slightly older medical cases, and the sort of more complicated and if you like sicker or frail your patients get, there’s a massive trade off then for what it’s possible for us to do and actually what we should be doing*.
*[ED53] Variability in response depends on type of pre-alert - we get a lot of calls where it’s a bit like “Geriatric patient with sepsis who’s 95” with a pre-alert. My brain automatically goes “Ok, it’s an unnecessary call…However, something which is rang through by Helimed - automatically picks up the ears*.
*[ED55]: I think it can have a negative effect if that phone’s constantly going, because you start getting sick of answering it. You can’t get on with anything, you’re frustrated, perhaps not gonna be listening as well…They don’t know that we’ve just had 5 phone calls or there that were courtesy calls and this is the 6th one, and you’re doing my head in! That’s not their fault*.

### 2. Individual clinician factors affected the ED response

ED staff frequently received no or limited training about how to answer pre-alert calls and there was evidence of variation in pre-alert response (i.e. where the patient would be brought) between different ED clinicians depending on the experience, attitudes to risk, situational awareness, and whether there were established relationships with ambulance clinicians. Information provided by ambulance clinicians was usually ‘accepted’ but decision-making also depended upon trust in information provided. We observed staff directing some pre-alerts to the ‘front door’, dismissing them as driven by risk-averse protocols.

*[Site A_MTC_Obs_3] Pre-alert 10. Consultant in charge looks over at what’s being written on the pre-alert form and laughs. “[condition recently added to ambulance service guidance for pre-alert]?”. Says send them to front door. Takes form to [Consultant] and [Nurse in charge]. [Consultant] comments that this is utterly insane*.
*[ED42]: Certainly if the senior paramedic’s brought the patient in and they’re saying they’re really poorly I’ll listen to them. I mean that’s just, if they’re concerned about the patient then I’m concerned as well*.
*[SiteA_MTC_Obs5a] Conversation with [role]: ‘Some crews alert everything, whereas others if they say someone is sick, you know they are sick, take it seriously whatever the obs.’*

EDs lacked formal policies for responding to calls, with most expressing an ‘understanding’ that the call will be answered by a senior clinician (nurse co-ordinator or senior doctor), but others reporting a ‘whoever is nearest’ policy or this being the reality in practice.

*[SiteE_MTC_Obs1:] We are told NIC [Nurse in charge] mostly answers but this wasn’t observed. Anyone registered can answer but usually the more confident. NIC says there is no set list of questions - “go off what the crews say”. Call sign noted first, always given by [ambulance service 3]*.
*[SiteA MTC Obs2]: Registrar took calls, other staff watched) - senior doctors or nurses said they answer red phone but SHO said “I pick it up all the time”*
*[ED23] Anyone can pick the phone up. Which most of the time is not anybody that picks it up, it’s usually one of the senior clinicians or senior nurses that pick it up. Because they know what to do with the information*.
*[ED12] We’re encouraged from fairly junior all the way through to senior to answer that phone. Particularly as a trainee I’m encouraged to answer that phone and hopefully try and see that patient then in resus if that’s suitable’*

This resulted in calls being taken by staff who were less able to respond appropriately to the call. When the pre-alert phone was answered by less experienced staff this created workload as they would often have to check where the patient should be taken with a senior clinician, or senior clinicians would get involved, directing the call taker to ask specific questions or checking that the correct advice was given.

*[SiteF_TU_Obs2:] F2 answers, more senior registrar nearby. The F2 reads out the details given - Choking episode. ‘I’m tempted to say that we wouldn’t accept this and go to [front door] but I’m not senior enough to say that so I’ll just check.’ checks with the more senior reg and say ‘go to [front door] initially’*.
*[SiteF_TU_Obs2]: F2 answers, NIC comes and stands behind. Consultant and more senior reg also there - unintentional overdose of codeine. The consultant circles GCS on the form to indicate what to ask. Consultant says he can go to [front door]*.
*[ED03]: If it’s really busy, junior doctor or junior sister/staff nurse will answer but they sometimes will forget to ask something even though it’s on the sheet so I think that’s why it is quite important that it is a senior member of the team or, say if I’m present, at least I’m stood next to them to kind of prompt them’ [ED03_Senior Nurse]*

Senior clinician response or involvement often led to improved processes and outcomes for pre-alerts, particularly for trauma calls.

*ED48: I think you need a clear identifier of who is going to be the person who picks up the pre-alert form and who is going to be the clinician that is involved…there needs to be a coordinating nurse and there needs to be a coordinating clinician*.
*ED15: think the nurse in charge should know and it’s not a control thing, it’s a safety thing. I think one person, essentially one person has that overview of the whole department. And if they don’t get fed that information then it can cause carnage*.

### 3. Processes for receiving, documenting and informing others of pre-alerts varied across EDs

Documentation processes varied in terms of the information collected and the flow of information following the call (see table 3). Pre-alert information was communicated to staff within the ED verbally (bleep or face-to-face) or via written information taken to the receiving area but information was not always consistently conveyed, particularly when patients were not brought into resus. Where processes for information flow following the pre-alert were less clear or were interrupted by other pressures this could lead to pre-alerts not being communicated appropriately, causing stress for both ED and ambulance clinicians.

*ED33: Whereas when sort of some of our junior doctors, they’ll just take the call, they won’t tell the crew where to go. And sometimes what they haven’t done, or done in the past, is just left the form on my desk and not told me about it, and we’ve had crews rock up with an alert and I don’t know about them*.
*ED15: Some doctors (especially junior) don’t clearly communicate pre-alerts or leave paperwork on desk, others will act on the information [take ownership!]*.
*ED36 Communication could be much better to make sure that everybody knows that an alert is coming in and because sometimes alerts arrive and nobody but the nurse in charge and the nurse in the area is expecting them. There are times when I don’t know anything about an alert and it will arrive in the department and or you’ll find that there’s an alert on an ambulance that we didn’t know about*.

Variation in information collated and inconsistent information flow following the pre-alert meant that there was usually minimal information that could be collated for audit purposes. Written information varied in quality and completion. A lack of patient identifiers (name, NHS number) on pre-alert forms at most sites, or information being discarded or remaining unattached to patient notes meant information was sometimes lost and therefore not usable for audit purposes.

*[SiteE MTC Obs3b]: NIC runs through the pre-alerts that morning. One for sepsis, NIC not sure if they’re in resus or in majors. Pre-alert form says [front door]. The patient has been sent to [front door] without pre-alert information*.
*[Role] informs us there is currently, no system for informing [front door] that the patient is a pre-alert not accepted in resus but it’s rare that that information wasn’t communicated. ‘If they’ve been told to go to [front door] then they would just take on the same process as all the other crews.’*
*[ED48]: I don’t think we do very well taking the pre-alert and going right, […], you’re gonna go straight to [front door] and then us physically taking that pre-alert to [front door]…can the nurse see them? And if they have any concerns, get a clinician or come to resus to get us to have a look at them.[…] My expectation is that somebody would take that piece of paper into [front door]. That doesn’t always happen*.

### 4. Opportunities for senior clinical review of pre-alerted patients who were not brought to resus varied between EDs

Potential pathways for pre-alerted patients differed across the sites, reflecting different approaches to managing risk, historical processes and capacity, layout and resourcing of the ED (see Table 3). These ranged from bringing the patient into resus or ‘high care’ areas, directing to the ‘front door’ for review by a member of staff upon arrival, or directing to the ‘front door’ to join the usual ambulance queue. Pre-alerted patients were usually (but not always) given higher priority. Within our fieldwork over one third of pre-alerts (53/143) were not told to go to resus or high care. Processes for assessing patients who were not brought into resus varied between sites. Whilst sites were often happy for pre-alerted patients to be sent to the ‘front door’, they sometimes undertook an immediate ‘eyeballing’ review of incoming pre-alerted patients as a check, partly in acknowledgement of the difficulties in assessing over the phone. One site also offered this review option for patients who had not been pre-alerted, meaning that ambulance clinicians could receive immediate reassurance about a patient they had concerns about but not pre-alerted.

*[SiteE MTC Obs3b]: Pregnant lady in resus not pre-alerted by double technician crew because ‘if it’s borderline we normally ring the doorbell and see what they want.’ Turn up and check has been observed at least three times now, maybe as ambulance door is close to resus so it’s easy to stop and check as the crew are walking through to [front door area]*.
*[ED48]: If I’m not sure, because you can’t really tell what a patient looks like from their numbers, if I’m not sure, then I’ll say, okay, that’s fine. Can I see them at the door? And I’ll make a decision based on physically casting eyes on the patient and physically speaking to them, and I think that is very, very useful*.
*[ED53]: If a pre-alert goes to [front door], it is given to the [front door] nurse and if it’s something that needs a quick review, the [front door] doctor will often be informed to prioritise it*.

### 5. Responding to pre-alerts involves significant staff time and resources

Responding to the pre-alert phone was always observed to be prioritised by staff, regardless of the level of demand at the time of call. The call also influenced the behaviour of other staff, who often listened in, read what was being written and sometimes started to act before the call was complete. The pre-alert provided an opportunity to prepare for the incoming patient’s arrival, with the response depending upon the severity and needs of the incoming patient. Resource use was particularly high for complex cases (e.g. major trauma) or where immediate life support was requested, involving the readying of equipment and calling specialist teams from elsewhere in the hospital.

*‘A pre-alert is not a harm free intervention. Sometimes in resus there is only 1 spare bed. And a pre-alert might take a reg, ED nurse, anaesthetist, consultant, [allied health professional].’ [SiteD MTC Obs3a]*
*It allows me to plan where the registrars go, which resus beds we’re going to use, who we’re going to use, who’s going to go in each bed, maybe we don’t start to do a sedation procedure which ties up staff for a long period of time. We know that there’s something coming in. So, I suppose the knowledge allows us to use the resources the best that we possibly can use them. [ED30_Consultant]*
*[Site A MTC Obs1] Consultant in charge talks about the impact of pre-alerts on the whole hospital, particularly when the one person on call in the whole hospital is in ED. “Nothing is going on elsewhere” - emergency theatre stops, ward rounds stop*.
*(Later that morning. Pre-alert 10:15. Transfer from trauma unit - complex major trauma. Results in many follow up calls and waiting to hear reasonable ETA)*
*10.57 Consultant passing info to nurse and doctor waiting by pre-alert phone – patient has set off, more info conveyed. Conversations between various members of staff. Move equipment and trolleys so there is room for people. Wheels ultrasound over*.
*11.04 Consultant pushing screens back for more space. Staff put on their role labels. More info is added to board. Call going out to trauma team & cardiothoracic. Sister checking notification has gone*.
*11:08 People start arriving – major trauma consultant, radiographer, others, discussing plan of action. Five staff in bay, another five by reception*.
*11.14 formal briefing. Now there are 17 staff in the area waiting. Going through basics, what needs doing, by who. Consultant asks - any questions? Checking re neurosurgeon, agree to call once patient here. People discussing roles. Consultant is discussing treatment plan. People being asked to sign in on checklist*
*11:24 patient arrives*

Due to the pressures in the department, creating space for incoming patients often involved ‘stepping down’ critically ill patients who would otherwise have remained in resus, whilst also trying to retain space for sudden changes. The response often involved “juggling available space and staff” (ED56), including requesting immediate portering to move patients and ensuring staffing levels were sufficient across all areas.

*[ED03] Having that information beforehand is really handy because if our resus room is full then we need to kind of step people down into the majors area or […] maybe move them further down the resus rooms so we’ve got an airway bay free*.
*[ED30]: It allows me to plan where the registrars go, which resus beds we’re going to use, who we’re going to use, who’s going to go in each bed, maybe we don’t start to do a sedation procedure which ties up staff for a long period of time. We know that there’s something coming in. So, I suppose the knowledge allows us to use the resources the best that we possibly can use them*.
*[ED36]: It does enable us to work out where people are going to go and start to try and move people around in advance and certainly for the sick patients that’s really important and that you know the longer warning we can get, the better in many ways*

Due to the workload involved in acting on pre-alerts, timing of the call was perceived as critical to maximising the benefit for both patients and staff. Notice of 10-15 minutes was generally considered optimal to enable necessary changes to be made, although more time was valuable when substantial preparation was needed. ED clinicians expressed frustration when timings were under-estimated, resulting in wasted resources, especially when large teams of specialist staff had been called. Short notice calls (less than 5 minutes) were also challenging, but ED staff valued any pre-warning even when crews were very close, and expressed frustration when ambulance clinicians did not do this, or left the call to the last minute.

*[ED33]: A lot of the time it’s to do with distance, so they’ll say well we were only five minutes around the corner, which is the most common reason to be honest. But it still would have given me four or five minutes to set things up for this patient, and that is part of their protocol so it’s a bit unacceptable really, but again I always speak to the crews*.
*[ED49]: I would rather a crew rung me and said, I’m two minutes away. I’ve got a really poorly one, can you look at them? So then at least I’ve got those two minutes to mentally say like, right, this patient can move here, this patient can move here*.
*[SiteA MTC Obs2]: Sister mentioned that a patient in resus that had not been pre-alerted as they were close to the hospital*.
*[SiteE MTC Obs2b]: 12.50 Critical care paramedics bring a patient in that wasn’t pre-alerted. Apparently this happens often. I asked them why they didn’t pre-alert and they said it was because they were only 2 minutes away*.
*13.10 [Speaking to trauma consultant] He told me his bugbear, which is wildly inaccurate ETAs. He said they can be waiting from 5 minutes to 1 hour. Yesterday there were 2 instances of waiting for pre-alerts, of waiting over half an hour and nearly an hour and the pre-alerts never turned up*.

### 6. High demand led to an increase in pre-alerts but also increased the importance of pre-alerts

Pre-alerts were perceived to be more important in the context of increased workload due to the increased risks from ambulance waits. When the systems were under increased pressure within limited space then pre-alerts enabled them to act, even at short timescales.

*[ED18]: I think the pre-alert is even more important now, because it makes the difference between knowing you’ve got to find a space that is gonna be hard to find for someone in an immediate timeframe, versus them being able to wait for twelve hours on the back of a truck*.
*[ED19]: At the moment with the departments across the country being so busy often there isn’t just a spare space that we can take someone who’s really sick, so at least it gives us, even if it’s just five minutes or sometimes they give us 20 minutes or half an hour, just to prepare some space, like physical space for the patient to come into*.
*[ED17]: You can see how very easily everything becomes very manic, because we haven’t got the space. So the more, the earlier we know about these sick people, the better, cos it just gives us a little bit of time to try and prepare*.

However, this additional pressure was also felt to lead to an increase in pre-alert calls for cases that were not necessarily patients in need of an immediate different clinical response, but where ambulance clinicians had concerns about the patient having to wait in the ambulance. This resulted in a higher volume of calls to the pre-alert phone, increasing workload and adding another layer into the process of ED triage.

*[ED46]: Now, we get a phone call where actually it’s not really a critically ill patient. It’s just for advice, you know “What do we do with…?” and this has come out of the fact that, if they can’t drop the patient off in resus, then we are committing them to queueing up in the corridor for hours. [ED46_Consultant]*
*[ED56]: I think now we get potentially more courtesy if you like, courtesy calls or can I let you know we want to bring this one, and if we don’t think they’re well enough to wait, if handover is delayed*.

Whilst acknowledging the validity of clinician concerns and potential patient safety benefits, individual clinician attitudes towards these calls and perceptions of what the ‘red phone’ should be used for caused some frustration and tensions due to the increase in workload created. Some perceived that advice calls should be managed elsewhere, potentially via an alternative line or ambulance service support.

*[ED44]: A lot of medical calls you’ll pick them up and it’ll be this is the situation, can I run it past you?, yeh that’s fine but you’ve got a clinical team in your control room which you should be using but it’s fine. [ED44_ Nurse]*
*[ED55]: I know we get quite a few where they may be more junior paramedics who will ring up for advice on the pre-alert phone to say y’know I’m not sure if this needs pre-alerting, but this is what I’ve got. Which of course, nobody minds that, it’s just difficult. If you’ve got a busy resus, and that phone’s constantly going, and they’re not pre-alerts. So it’s almost like we need a pre-alert phone and an advice phone*.
*[Site D Obs3a]: Another thing that bugs me is when they just call for advice. This is an information line not a discussion line. [Ambulance Service 2] have a trauma desk if they want trauma discussion*.

### 7. ED clinicians valued pre-alerts and perceived higher risks from under-alerting than over-alerting

However, overall ED clinicians described a risk-averse approach to pre-alerting, perceiving “it’s better knowing about them than not knowing about them” (ED4). They also reported concerns about certain conditions being under-alerted (e.g. silver trauma). Being given information about borderline cases enabled them to prepare for potential deterioration and the chance to ‘eyeball’ a patient if necessary.

*[ED30]: If you’re being pragmatic about it, any information is useful. And I think that just because they’re pre-alerted doesn’t necessarily mean we have to respond in a way that they would perceive they would want to be responded to*.
*[ED48]: I personally think that we should all at least see them very briefly to just clap eyes on them. That’s a respect thing for saying that, you know what, no, I’m not worried about them, but actually thank you for calling*.
*[ED30]: I’d rather have all of the information and then make the decision when they arrive, nothing is ever dismissed.’*

Pre-alert communication and judgement was sometimes inaccurate but the consequences of under-alerting were felt to be more serious than those of over-alerting, putting staff under extra pressure when they have not had chance to prepare. Even short notice pre-alerts enabled some mental preparedness through awareness of the risk profile of patients arriving into the ED.

*[ED49] I would rather a crew rung me and said, I’m two minutes away. I’ve got a really poorly one, can you look at them? So then at least I’ve got those two minutes to mentally say like, right, this patient can move here, this patient can move here*.
*[ED42]: We sometimes get people who are just brought in who aren’t alerted, sometimes from high speed accidents but they don’t trigger on necessarily the pre-hospital trauma tool, but they would trigger on our tool because it’s slightly different. And actually, I’d much rather be alerted and then have to step down.’*
*[ED38]: In any triage system you have to overtriage, if every call you’re bringing in is entirely appropriate then you’re missing things…Anything that encourages people to alert less, it’s fraught with danger*.

Even when staff did not make any immediate practical change within the department in response to the pre-alert, they still valued the pre-alert as enabling them to mentally plan and prepare, giving them some sense of agency in a chaotic, unpredictable environment.

*[ED18]: Part of the whole essence of pre-alerts is for your own mental model, isn’t it? It’s for your own preparation*.
*[ED25]: I don’t find that we get pre-alerted too many things, I find them nearly all helpful*.
*[ED43]: You are aware of what’s there, it’s not a hidden risk, it’s a, you’re aware of the risks*.

## Discussion

We identified that EDs had different processes and practices for managing pre-hospital pre-alerts that resulted in different responses to calls. A complex interplay of factors impacted on the response including resource availability within the department at the time of the call, but also consideration of how cases could be managed elsewhere, risk perception and individual clinician practice. Pre-alert calls were taken seriously and resulted in significant work for EDs, which amplified the importance of accurate estimated arrival times and high quality information handover. Despite some risk of pre-alert fatigue for certain conditions (e.g. sepsis) pre-alerts enabled staff to plan both practically and psychologically for a patient’s arrival and to manage wider patient flow within the department. In general, pre-alerts were highly valued, with the risks of under-alerting perceived as worse than those of over-alerting. EDs had different processes to enable review of ambulance patients, both those pre-alerted and not, which may impact on ambulance clinician pre-alerting behaviour.

We identified limited research exploring the use of pre-alerts beyond the literature identifying improved outcomes for specific patient groups who are pre-alerted. (2-7), (22) Our findings identified that EDs respond to a wide range of pre-alerts outside of these conditions by preparing space, personnel and equipment as highlighted within the literature, but also prepare psychologically for the patient’s arrival which is key to ensuring safe patient flow. We identified that pre-alerts did not always result in a resus bed, particularly when resources were pressured, but that EDs frequently responded in other ways which increased their priority and helped staff to manage associated excess risk. These processes included ‘eyeballing’ them on arrival, or putting them in a higher care area. Sujan et al identified pre-alerts had an important anticipatory function in enabling EDs to prepare for the patient’s arrival.(18)

ED crowding and prolonged waiting times are associated with increased mortality and a negative impact on other patient outcomes. (23) Within this fieldwork EDs were frequently crowded, operating beyond capacity with long ambulance queues and were often unable to create space in resus for patients who would otherwise have been considered to warrant a resus bed. Pre-alerts also resulted in critically ill patients being moved (stepped down) in anticipation of a higher acuity pre-alerted patient. The ED response was more nuanced than ‘accepted’ into resus or not; patients were still usually considered higher risk within the ED clinician mental model of patients within the department and managed accordingly. The complex interplay of factors affecting decisions meant the ED’s response to similar patients was not always the same, which could cause confusion for ambulance clinicians.

Although we did identify some concerns about ‘pre-alert fatigue’ due to over-alerting (notably in relation to sepsis), this did not largely appear to affect ED clinician’s behaviour, and pre-alerts were generally taken seriously and prioritised. Over-alerting for sepsis may reflect the poor predictive value of diagnostic impression and early warning scores for sepsis and limitations of the sepsis diagnostic definition in a typical ED population. (24) Over-alerting did not appear to generate significant risks to other patients because ED clinicians made their own assessment of the patients needs and provided a graduated response, only freeing up space in resus when it was safe and necessary to do so. Berglund et al 2012 identified that stroke prenotification improved time to thrombolysis, with no negative impact on other prehospital patients. (25) Brown and Bleetman identified under-alerting to be a greater problem than over-alerting in a small sample of 52 critically ill patients, of whom 29 were not alerted. (26) They presented an ideal model of pre-alerting that included all critically ill patients, plus some non-critical patients to allow for a ‘margin of error’. Sheppard et al identified a lower proportion of patients were not pre-alerted for stroke who should have been than those who were pre-alerted who shouldn’t have been. (17)

Pre-alerts were shown to take up staff time and generate interruptions to caring for patients already in the department. Although we did not quantify time ‘wasted’, unnecessary interruptions and inappropriate pre-alerts were a source of frustration to ED staff. Despite this, overall pre-alert calls were usually perceived to be beneficial. Interruptions are common in the ED and can impact negatively on patient safety. However, interruptions may also be beneficial when providing critical time-sensitive information relating directly to patient care, as is seen with pre-alerts. (27, 28)

### Limitations

Although we aimed to represent a diverse population, there may be limitations to the transferability of findings. The three ambulance services primarily followed a ‘direct to ED’ call model whereas some other ambulance services use a call desk model. Half of our sample were major trauma centres, which were perceived by ambulance clinicians in our fieldwork to manage pre-alerts better than local EDs where pre-alerts are less frequent. Observations were undertaken at times that EDs reported most pre-alerts occurred. This meant we undertook few observations at night or weekends, when staffing levels and balance may differ. It is also possible that some behaviour modification took place as a result of our presence as observers. Although we reached saturation during observation and interviews for the main themes, we lacked data to explore certain themes (e.g. role and seniority) further.

### Implications of the results for practice or policy

Whilst a considerable level of variation in response to pre-alert calls is outside the control of ED staff, particularly in the current context of high demand, there is potential to increase consistency in some areas. Simple guidance and training could help EDs review and clarify their practice in relation to who answers pre-alert calls and how; who makes decisions and how; what information is documented; how information regarding the pre-alert is communicated to others, including when they are not accepted into resus. There is also a need to ensure decisions and protocols are disseminated to all staff, potentially through brief training or other mechanisms. This is particularly the case given the rapid turnover and frequent rotation of staff within EDs.

There is a need for further alignment of ED and ambulance service policies and pre-alert thresholds for some conditions, particularly sepsis, and for identifying routes for ambulance clinicians to seek advice on patients they are uncertain about. There is also a need to increase ambulance service awareness of the complexity of ED decision-making regarding pre-alerts to avoid misunderstanding and tension when ambulance clinicians do not receive an anticipated or consistent response which could impact negatively on future pre-alert behaviour.

## Declarations

## Ethical considerations

Ethical approval for the pre-alerts project has been obtained from Newcastle & North Tyneside 2 Research Ethics Committee (Ref: 21/NE/0132)

## Authors’ contributions

JL organised and undertook fieldwork, led data analysis and contributed to the writing of the paper. FS conceived and designed the study, contributed to the analysis and drafted the paper. JC contributed to the study design, oversaw PPI involvement, undertook fieldwork and contributed to the analysis and writing of the paper. ROH contributed to the study design, undertook coding and analysis and contributed to the writing of the paper. FB contributed to the study design, contributed to analysis and interpretation of the data, critically revised the paper and approved it for final version. SG contributed to study design, contributed to interpretation of the data, critically revised the paper and approved it for final version. All authors read drafts of the manuscript and approved the final version. JL is guarantor for the paper.

## Data Availability

The data generated for this study is in the form of confidential transcripts of interviews that are not available for sharing. Participants consented for anonymised quotations to be shared but did not consent to share the full transcripts.

## Acknowledgements

The authors would like to thank all research participants and ED and ambulance service staff who helped to recruit participants. We are also grateful for the input of other members of the study team, our advisory group and our patient and public involvement representatives/group. Thanks to Marc Chattle for clerical support.

## Patient and Public involvement

Patient and public involvement (PPI) representatives contributed to the design and conduct of this study through input at PPI and project management group meetings. Preliminary findings were presented and discussed at a PPI event for the study.

